# An EMG-Based Implementation of the Distal Upper Extremity Tool: A Proof-of-Concept Comparative Evaluation

**DOI:** 10.1101/2025.10.03.25337266

**Authors:** Xuelong Fan, Johan Rydgård, Pasan Hettiarachchi, Kristina Eliasson, Camilla Dahlqvist, Peter J. Johansson

**Author notes:** Corresponding author: Xuelong Fan.

## Abstract

Work-related distal upper extremity (DUE) disorders are common in hand-intensive occupations due to cumulative strain. Existing assessment tools rely on subjective ratings or summary exposure metrics that assume homogeneous work patterns, limiting their ability to capture time-varying loading across tasks and schedules. This proof-of-concept study evaluated whether an electromyography (EMG)-based implementation of the Distal Upper Extremity Tool (EMG-DUET), combining sensor-based exertion estimates with cumulative damage modelling, could provide DUE symptom risk rankings. Nineteen participants were video recorded at work with forearm EMG, yielding 78 tasks. EMG-DUET was compared with observation-based DUET (OBS-DUET), forearm EMG peak values (EMG-Peak), the ACGIH® Hand Activity Level Threshold Limit Value (OBS-HATLV), and occupational DUE symptom prevalence estimates. Both DUET variants aligned most closely with benchmark tools using the same input modality. EMG-based tools showed numerically stronger, though non-significant, correlations with prevalence than observation-based tools. Therefore, EMG-DUET remains exploratory and requires refinement and prospective real-world validation.

## 1 Introduction

Work-related distal upper extremity (DUE) disorders remain a major concern in occupational health (Govaerts et al., 2021). Physical workload, particularly in hand-intensive tasks, plays a key role in DUE disorders such as epicondylitis and carpal tunnel syndrome, primarily through cumulative tissue damage and chronic inflammation (Dong et al., 2022; Keir et al., 2021).

Effective workload assessment is critical for predicting and managing the risk of work-related DUE disorders. Existing tools differ along two important dimensions: how physical exertion is measured and how exposure is integrated across tasks and time.

The first dimension concerns the source of exertion information. Self-reported and observation-based approaches rely on ratings of exertion or work activity (Grant et al., 1999; Kapellusch et al., 2014; McAtamney & Nigel Corlett, 1993; Robertson et al., 2003; Yung et al., 2019). Self-reported measures may be affected by recall bias, workflow disruption, and changes in perception or behavior during momentary reporting (Bell et al., 2019; Ortega et al., 2024). Observation-based tools, including the ACGIH® Hand Activity Threshold Limit Value (OBS-HATLV) (Yung et al., 2019), can be applied through on-site or post-hoc video assessment with minimal disruption to workers. However, they remain susceptible to inter-observer variability and systematic bias, including sex bias, while manual evaluation makes repeated or long-term assessment resource-intensive (Dahlgren et al., 2022; Graben et al., 2022; Nyman et al., 2023). In contrast, technical measurements such as surface electromyography (sEMG) provide continuous, sensor-based measures of muscular activity. sEMG-derived measures of forearm muscular load have been associated with DUE disorder risk and are commonly interpreted using empirically derived action levels (Arvidsson et al., 2021; Balogh et al., 2019; Nordander et al., 2013).

The second dimension concerns how exposure is represented across tasks and time. Many conventional tools characterize a task or job using a peak, average, or representative exposure value over a predefined assessment period (Arvidsson et al., 2021; Yung et al., 2019). This approach implicitly assumes that the assessed work pattern adequately represents the wider exposure period and is therefore best suited to relatively homogeneous work. In practice, however, workers may alternate among tasks with different exertion intensities, repetition rates, durations, and recovery opportunities, both within and across workdays (Bolino et al., 2021). Summary measures may therefore fail to preserve the time-varying loading history of task-diverse work. From a fatigue-failure perspective, cumulative damage depends on the distribution of repetitions across exertion levels, requiring exposures to be integrated across tasks rather than represented by a single summary value (Gallagher & Schall Jr., 2017).

The Distal Upper Extremity Tool (DUET) addresses this temporal-modelling limitation by estimating cumulative risk across multiple hand-intensive tasks. Its framework applies material fatigue-failure theory to estimate cumulative tissue damage from exertion intensity and repetition, and then maps this damage to the probability of DUE symptom occurrence using an epidemiological model (Gallagher et al., 2018; Mehdizadeh et al., 2020). By calculating damage at the task level, DUET can integrate exposures across user-defined periods, multiple tasks, workers, and job rotations (Jorgensen et al., 2024; Mehdizadeh et al., 2020).

Neither approach alone, however, addresses both dimensions. Conventional EMG-based measures, such as peak forearm muscular load (EMG-Peak), provide sensor-based exposure information but reduce the recorded loading history to a summary metric. Conversely, the original observation-based DUET (OBS-DUET) integrates exposures across tasks but retains rating-based exertion inputs. An EMG-based implementation of DUET (EMG-DUET) could combine the complementary advantages of both approaches: sensor-based exertion estimates from sEMG and cumulative modelling of task-varying exposure through the DUET framework. However, EMG-derived exertion estimates have not previously been integrated into DUET. In the present proof-of-concept study, this integration was evaluated at the task level as a necessary first step toward cumulative assessment across real work schedules.

Together, EMG-DUET, OBS-DUET, EMG-Peak, and OBS-HATLV enable the two dimensions to be evaluated separately: input modality by comparing EMG-based and observation-based tools within the same modelling framework, and modelling framework by comparing DUET and non-DUET tools within the same input modality.

Therefore, as a proof of concept, this study explored whether EMG-DUET could provide meaningful risk rankings for DUE symptoms in hand-intensive work. We evaluated its risk rankings against OBS-DUET, the non-DUET benchmarks EMG-Peak and OBS-HATLV, and occupational-level prevalence estimates of work-related DUE symptoms (Statistikdatabasen, 2023).

## 2 Materials and methods

### 2.1 Participants

Nineteen participants were recruited for the study, of whom 26% were female. The mean age was 37 years, and the average body mass index (BMI) was 26 kg/m². Most participants (84%) were right-handed. Task assessments were performed on each participant’s preferred side, except for one individual who was assessed on both sides. For those evaluated on the left side (N = 3), the average maximal left-hand grip strength was 36 kg, while for right-sided assessments (N = 17), it was 50 kg. (**Table 1**). All participants provided written informed consent prior to participation, and the study was approved by the Swedish Ethical Review Authority in Gothenburg (Dnr 2022-02531-01).

**Table 1.**
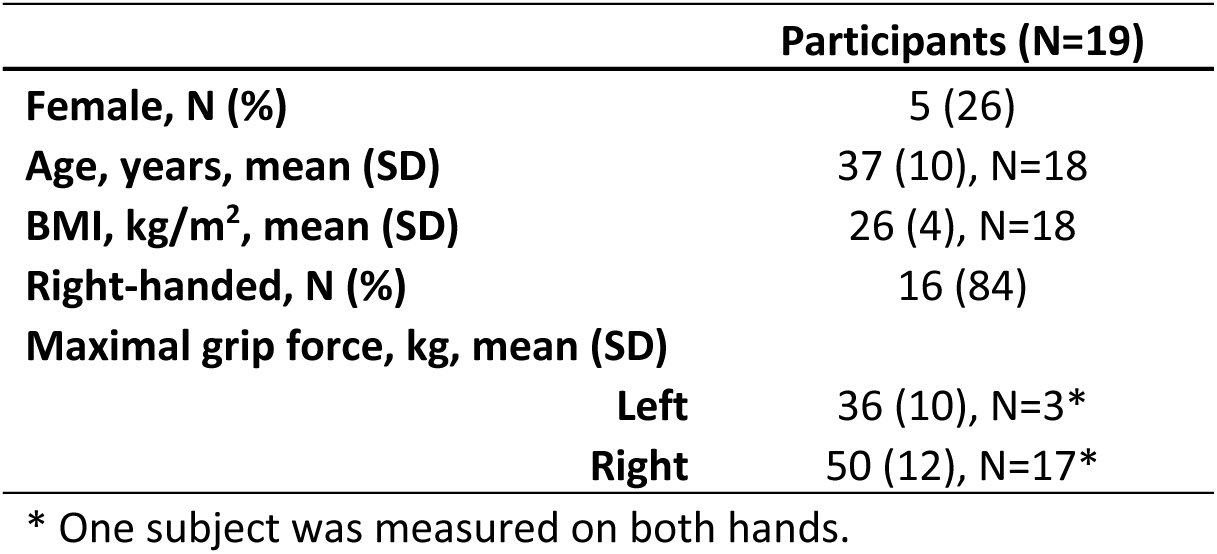
Participant demographics (N = 19). For variables with missing data, the number of participants with available information is indicated by N. If not specified, data were available for all 19 participants. SD stands for standard deviation.

### 2.2 Tasks

Seventy-eight DUE-related tasks were recorded between 2022 and 2024. Tasks were selected by convenience sampling to maximize diversity across task types and occupational settings, and represented typical work activities from 14 occupations and industries in Sweden. Task selection was guided by professional ergonomists, participating companies, and workers. Each task was labeled by occupation abbreviation, occupation number, participant ID, and assessed hand side (Error! Reference source not found.). Participants could perform multiple tasks; all tasks from a given participant came from the same industry or occupational category, except for M04, who performed tasks from two categories. Mean task duration was 7.3 minutes, with an interquartile range of 3.9 to 9.5 minutes.

### 2.3 Protocols

The measurement protocol consisted of two phases: a **calibration phase** and a t**ask phase**. Data collection was performed at the participants’ workplaces.

#### 2.3.1 Calibration phase

Calibration was done in the same way as the previous study (Fan et al., 2026). Participants were seated with the working arm positioned 90° from the trunk and the wrist neutral. They then held a suspended hand dynamometer (G200, Biometrics Ltd, Newport, UK), rested for at least one minute, and performed a maximal voluntary contraction (MVC) by squeezing the handle as forcefully as possible for at least two seconds, with verbal encouragement. Muscle activation was recorded using sEMG throughout the MVC.

#### 2.3.2 Task phase

In the task phase, participants carried out one or more predetermined tasks, with the order of tasks being arbitrary. Each participant was instructed to perform their tasks as naturally as possible in their usual workplace environment, verbally reporting their perceived exertion for each self-defined effort. A revised OMNI-Resistance Exercise Scale (OMNI-RES), adapted specifically to forearm exertion and ranging from 0 (no effort) to 10 (maximal effort), was used (Fan et al., 2026; Rydgård, 2025a). Throughout the tasks, muscle activity was continuously monitored via sEMG, and video footage was recorded from one or two camera angles (GoPro, Inc., California, USA) at 25 frames per second.

### 2.4 Muscle Activity

Forearm muscle activity was recorded using a through-forearm placement of sEMG electrodes. Through-forearm sEMG is a simplified electrode placement that captures a composite signal from the forearm flexor–extensor region and was used to assess exertions during hand-intensive work because it captures a composite signal from the forearm flexor–extensor region with a simplified electrode placement (Fan et al., 2026; Takala & Toivonen, 2013). The recording device and placements of electrodes were the same as the previous study (Fan et al., 2026). Two self-adhesive bipolar Ag/AgCl electrodes (N-00-S/25, Ambu, Penang, Malaysia) were applied: one over the belly of the m. extensor digitorum communis and the other over the m. flexor digitorum superficialis. A ground electrode was placed on the olecranon process at the elbow. Raw sEMG signals were captured with a digital data logger (DataLog MWX8, Biometrics Ltd., Newport, UK) and sampled at 1000 Hz per channel via a 24-bit A/D converter embedded in the logger.

### 2.5 Observation-based exertion

After data collection, an experienced ergonomist reviewed the video recordings of each task to identify individual repetitions and assess their intensity. Intensity ratings were assigned using the revised OMNI-RES scale, based on both the ergonomist’s expertise and, when available, participants’ self-reported exertion ratings. For static exertions lasting longer than 5 seconds, the number of repetitions was calculated by assigning one additional repetition for each subsequent 5-second interval (for example, 5–10 seconds = 2 repetitions, 10–15 seconds = 3 repetitions, etc.). The analysis was performed using a custom annotation tool, ErgoAnnotation (Rydgård, 2025b), integrated into Blender (version 4.4.3; Blender Foundation, Amsterdam, Netherlands).

### 2.6 Data processing

Raw sEMG signals were processed using an established pipeline that included filtering, smoothing, denoising, and normalization in preparation for exertion prediction (Fan et al., 2026). Initially, raw sEMG data were filtered with a digital bandpass filter (20–400 Hz) and an additional 50 Hz notch comb filter (Q factor = 35) to eliminate powerline interference and other high-frequency artifacts, resulting in the base sEMG data. The base sEMG signals were then smoothed using a 0.125-second root mean square (RMS) moving window. To account for background activity, the smoothed signals were power-subtracted by the resting noise level. This resting noise level was estimated as the minimum value of the base sEMG during the resting period, further smoothed by a 2.375-second RMS moving window. The power-subtracted signal was calculated as follows: 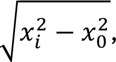 where xᵢ is the base sEMG value at time point i, and x₀ is the resting noise level.

Finally, the denoised and smoothed signals were normalized to the participant’s maximal voluntary electrical activation (MVE), expressing muscle activity as a percentage of MVE (%MVE). MVE was determined by the maximum value of the base sEMG signal during the MVC period, smoothed using a 0.5-second RMS window and power-subtracted using the same resting noise level. The resulting normalized signal (%MVE) was then used in all subsequent analyses.

### 2.7 Estimation of exertion

Observation-based exertion ratings were used directly as observational input. EMG-based exertion was estimated from normalized muscle activity using a previously established model (Fan et al., 2026), which converted sEMG (%MVE) into estimated exertion (%MVC) as described in **Eq. 1**:

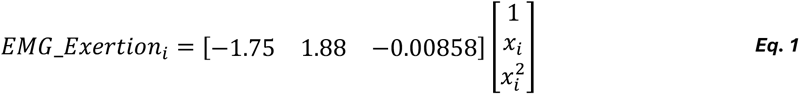

where *x*_*i*_ is the normalized muscle activity (%MVE) at the time point *i*, and *EMG*_*Exertion*_*i*_is the estimated exertion (%MVC) from EMG at the same time point.

Notably, the EMG-to-exertion conversion was used as a pragmatic approximation for this proof-of-concept implementation. It was not assumed to provide universally valid force estimates across all postures, contraction types, or tasks.

### 2.8 Risk assessment outcomes

In total, four risk assessment tools were applied to all tasks to estimate risk outcomes for the DUE symptoms, including 1) EMG-DUET and 2) OBS-DUET, and two other non-DUET-framework benchmarks, 3) EMG-Peak and 4) OBS-HATLV.

#### 2.8.1 EMG-based DUET (EMG-DUET)

EMG-DUET integrated EMG data into the DUET framework by using EMG-based estimates of exertion as input to predict the probability of developing DUE symptoms (Gallagher et al., 2018). Continuous exertion estimates from EMG readings were processed with a Rainflow algorithm to classify the intensity and frequency of distinct exertion cycles for each task (Nail-Ulloa et al., 2025). The fatigue-failure model within the DUET framework estimated tendon strain from exertion intensity, combining this with cycle frequency to calculate cumulative tissue damage based on theoretical cycles to failure, as derived from cadaveric research (Schechtman & Bader, 2002). Cumulative damage was then normalized to a cumulative damage density (per unit time) according to task duration and projected to a 6-hour continuous work exposure, enabling comparison across different tools. Finally, the calculated cumulative damage was mapped to the probability of DUE symptom occurrence using the DUET framework’s epidemiological model.

To ensure consistency between tools, EMG-based exertion values below 5 %MVC—the minimum exertion considered in OBS-DUET (corresponding to OMNI-RES = 0)—and exertion duration less than 0.25 seconds were excluded, as 0-rated exertions and exertions less than 0.25 seconds were not annotated in OBS-DUET.

#### 2.8.2 Original DUET (OBS-DUET)

OBS-DUET followed the original DUET framework and the same analytical pipeline as EMG-DUET, but used observation-based exertion ratings as input. All subsequent steps, i.e., strain estimation, cumulative damage calculation, normalization, and symptom probability estimation, were conducted identically to EMG-DUET.

#### 2.8.3 EMG-Peak

EMG-Peak was defined as the 90th percentile of through-forearm muscle activity (%MVE) (Fan et al., 2026). This differs from the proposed action level for peak forearm muscular load, which used extensor-specific EMG (Arvidsson et al., 2021; Nordander et al., 2004). Because no established action level exists for through-forearm EMG, the 30% MVE threshold was used as an approximate benchmark for this proof-of-concept comparison, not as a validated threshold for this electrode configuration. For this study, EMG-Peak refers specifically to the through-forearm measurement.

#### 2.8.4 OBS-HATLV

OBS-HATLV refers to a modified version of the ACGIH® HATLV® (Yung et al., 2019), which estimates the risk of carpal tunnel syndrome by combining measures of applied hand force and hand exertion repetition. To determine applied hand force, OMNI-RES scores for each exertion were first converted to Borg CR-10 using a matching table developed from empirical research (currently unpublished; **Table 2**). These Borg CR-10 scores were subsequently used to calculate the normalized peak force (NPF; Yung et al., 2019). HAL values were obtained by assessing repetition data from the annotation tool and applying a validated method (Radwin et al., 2015), then averaging across the task. Finally, a continuous risk score was calculated using Equation 2 (**Eq. 2**):

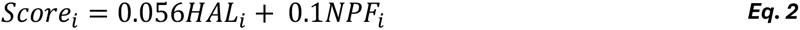

**Table 2.**
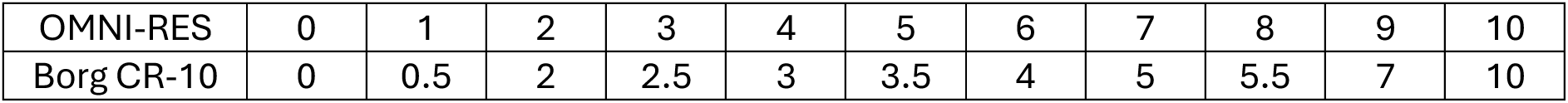
The matching table between OMNI-RES and Borg CR-10.

where *HAL*_*i*_ and *NPF*_*i*_ represent the estimated hand activity level and normalized peak force, respectively, for exertion *i*. Tasks were categorized as follows (Yung et al., 2019):

- Score > 0.56: High risk (above the Threshold Limit Value, TLV)
- 0.36 < Score ≤ 0.56: Moderate risk (between the Action Limit, AL, and TLV)
- Score ≤ 0.36: Low risk (below AL)

### 2.9 Prevalence of Work-related DUE Symptoms

The potential correspondence between tool-derived risk rankings and occupational health outcomes was explored using occupational-level prevalence estimates of work-related DUE symptoms. These prevalence rates served as proxies for DUE disorders across occupational titles. Data were sourced from national surveys conducted by Statistics Sweden (Statistikdatabasen, 2023), representing the average percentage of workers—regardless of sex—who reported work-related symptoms in the fingers, hands, or wrists during 2018, 2020, and 2022. Each occupation in the study was matched to its official title and code following the national classification system (Statistikdatabasen, 2014). Only occupations with at least one available prevalence data point from the surveyed years were included. In certain cases, multiple occupations shared the same prevalence value due to data being available only at a higher level in the occupational classification hierarchy.

### 2.10 Statistics

Analyses examined the tools along two dimensions, that is, input modality and modelling framework, at two analytical levels. At the task level, input modality was examined by comparing EMG-DUET with OBS-DUET, with EMG-Peak versus OBS-HATLV providing the corresponding comparison between EMG-based and observation-based non-DUET tools. Modelling framework was examined by comparing EMG-DUET with EMG-Peak and OBS-DUET with OBS-HATLV, thereby contrasting DUET and non-DUET approaches within each input modality. At the occupational level, the same two dimensions were explored by comparing the correlations with occupational-level DUE symptom prevalence between EMG-based and observation-based tools and between DUET and corresponding non-DUET tools. Three analytical approaches were used: risk-rank comparisons, risk-level classification comparisons, and outcome-rank comparisons. Analyses were conducted using MATLAB 2023a (The MathWorks, Inc., Massachusetts, USA).

Risk-rank comparisons were performed visually and quantitatively. Horizontal bar plots showed task rankings based on each tool’s risk scores, sorted by rank from the two benchmark tools (OBS-HATLV and EMG-Peak). Quantitatively, repeated-measures correlations (r_rm_) were calculated between tool-pair risk ranks to account for unbalanced repeated tasks per subject (Bakdash & Marusich, 2017). Ranks were subject-centered (value minus that subject’s mean), and Pearson’s correlation was computed on centered values for each tool pair. The 95% confidence intervals (CIs) were obtained using Fisher’s z transformation and back-transformed to r_rm_ (Silver & Dunlap, 1987). Statistical significance was set at p < 0.05. Correlation strengths were interpreted using established thresholds: negligible (<0.10), weak (0.10–0.39), moderate (0.40–0.69), strong (0.70–0.89), and very strong (≥0.90) (Schober et al., 2018). Because these thresholds are broad rules of thumb, correlations within the same category were also interpreted by relative magnitude when comparing tools.

Because no established risk thresholds exist for DUET tools, ROC analysis was used to evaluate how EMG-DUET and OBS-DUET classified risk relative to EMG-Peak and OBS-HATLV (Fawcett, 2006). For EMG-Peak, 30% MVE was used as the criterion; for OBS-HATLV, TLV and AL were used as criteria. Continuous benchmark scores were converted to binary high- and low-risk categories. ROC curves were generated across possible DUET thresholds, and AUC was used to quantify classification accuracy. Optimal cutoffs were selected using Youden’s J statistic (Youden, 1950), and sensitivity and specificity were reported at these thresholds.

For outcome-rank comparisons, task-level risk outcomes from each tool were averaged within participants and then aggregated at the occupational level to align with DUE symptom prevalence estimates. Spearman’s correlation (ρ) was calculated between each tool’s aggregated risk outcomes and occupational-level prevalence. The resulting correlations were then compared descriptively along the same two dimensions: between EMG-based and observation-based tools to examine input modality, and between DUET and corresponding non-DUET tools to examine modelling framework. Correlation strengths were interpreted using the same thresholds, with statistical significance set at p < 0.05.

## 3 Results

### 3.1 Risk-rank comparisons

Overall, the two DUETs displayed distinct characteristics in task-level risk assessment. EMG-DUET yielded a narrower range of risk probabilities, spanning from 58% to 80% (**Figure 1a**), while OBS-DUET displayed a broader distribution, with probabilities ranging from 18% to 90% (**Figure 1b**).

**Figure 1.**
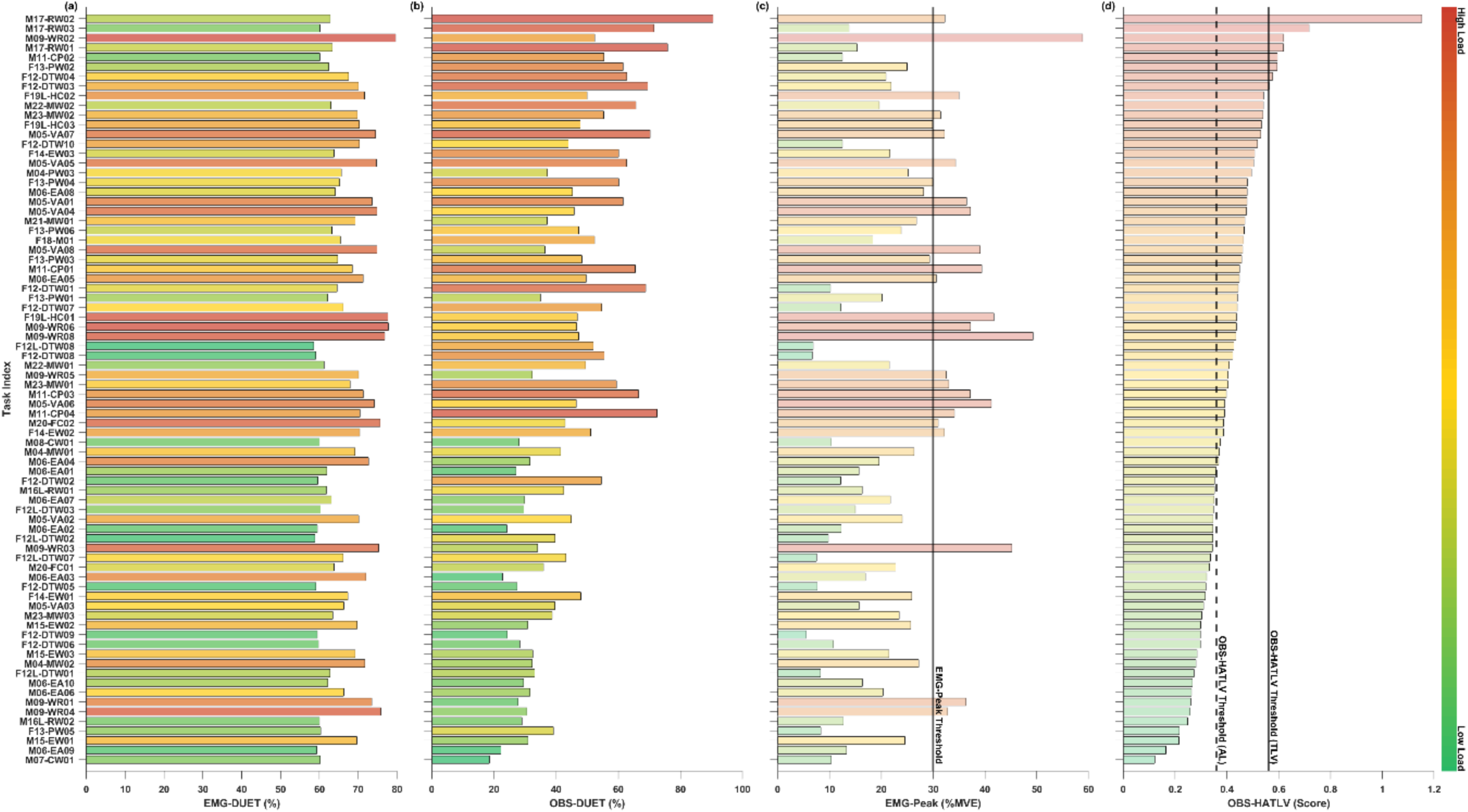
Comparison of risk assessment outcomes of tasks across (a) EMG-DUET, (b) OBS-DUET, (c) EMG-Peak, and (d) OBS-HATLV. Task index can be found in **Error! Reference source not found.**. Blacklines indicate the thresholds for categorizing risky tasks in the corresponding reference tools. Colors encode each task’s risk outcomes, ranging from low (cool hues) to high (warm hues).

Comparing the two non-DUET benchmark tools, differences emerged in their task classification. OBS-HATLV identified 8 tasks (10% of all tasks) as exceeding the TLV threshold and 39 tasks (50%) as falling between the AL and TLV thresholds (**Figure 1d**). By contrast, EMG-Peak, with values ranging from 6 %MVE to 59 %MVE, classified 24 tasks (31%) as having peak muscular loads above the 30 %MVE threshold (**Figure 1c**).

Significant correlations were observed between the DUET tools and their respective reference risk assessment instruments. EMG-DUET showed a significant positive moderate correlation with EMG-Peak %MVE, with a value closer to the strong than the weak threshold (r_rm_ [CIs] = 0.63 [0.45, 0.76], p < 0.001; **Figure 2c**), while the correlations with OBS-DUET (r_rm_ = 0.42 [0.18, 0.60], p < 0.001; **Figure 2a**) and OBS-HATLV (r_rm_ = 0.32 [0.07, 0.53], p = 0.014; **Figure 2d**) were weaker but still statistically significant. In comparison, the correlation between OBS-DUET and OBS-HATLV was strong (r_rm_ = 0.72 [0.57, 0.82], p < 0.001; **Figure 2f**), whereas the correlation between OBS-DUET and EMG-Peak, though also moderate, was more dispersed (r_rm_ = 0.45 [0.22, 0.63], p < 0.001; **Figure 2e**). As a reference, EMG-Peak and OBS-HATLV showed a significant moderate correlation, although the value was closer to the weak than the strong threshold (r_rm_ = 0.41 [0.18, 0.60], p = 0.001; **Figure 2b**).

**Figure 2.**
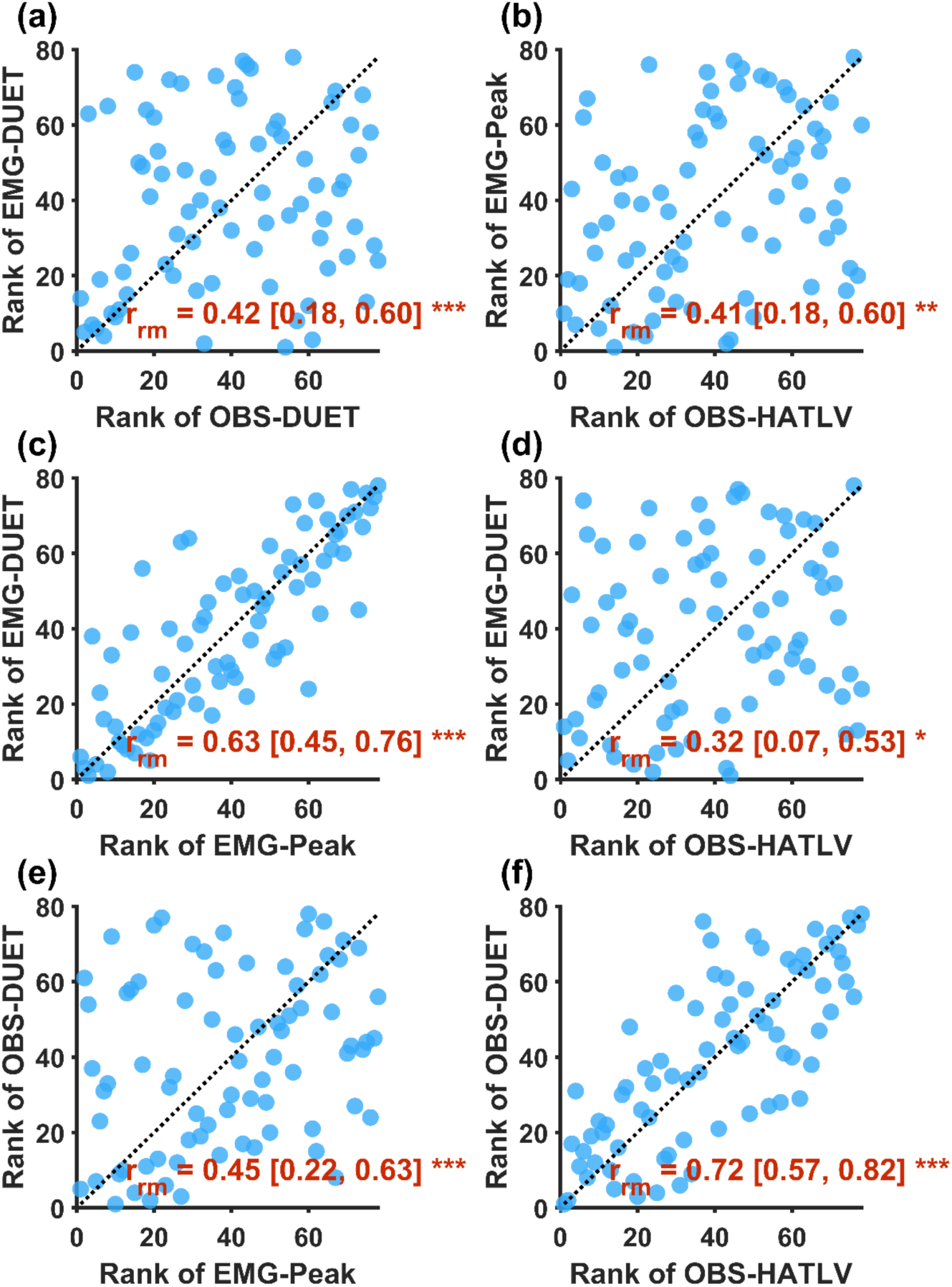
Rank–rank comparisons among DUET and reference risk assessment tools. Each blue circle represents a task’s rank in paired comparisons of EMG-DUET (y-axis) versus (a) OBS-DUET, (c) EMG-Peak, and (d) OBS-HATLV (x-axis); OBS-DUET (y-axis) versus (e) EMG-Peak and (f) OBS-HATLV (x-axis); and EMG-Peak (y-axis) versus OBS-HATLV (x-axis) in (b). The black dotted line indicates identity (y = x). Each panel reports the repeated-measures correlation coefficient (rrm) and corresponding significance: *, p<0.05; **, p<0.01; ***, p<0.001.

### 3.2 Comparisons of risk-level classification

When evaluated against the EMG-Peak reference (**Figure 3a**), the EMG-DUET exhibited high predictive accuracy, achieving an area under the curve (AUC) of 0.94. In contrast, the OBS-DUET showed moderate performance with an AUC of 0.68.

**Figure 3.**
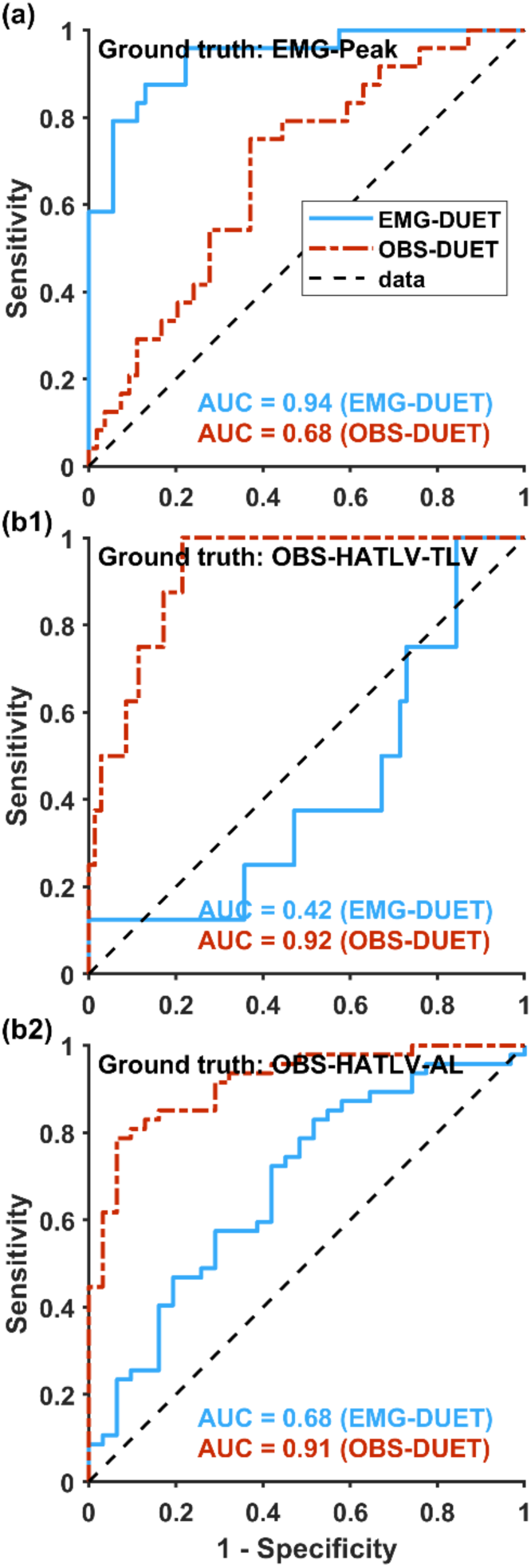
Receiver-operating characteristic (ROC) curves for EMG-DUET (blue) and OBS-DUET (red) risk scores against the two reference tools: (a) EMG-Peak, and (b1) the Threshold Limit Value (TLV) and (b2) the Action Level (AL) in OBS-HATLV. The black dashed line in each panel denotes chance performance (AUC = 0.50), and the inset AUC values report each DUET’s ability to distinguish between risky and non-risky tasks against the corresponding reference tool.

When compared to the TLV threshold in OBS-HATLV (**Figure 3b1**), OBS-DUET demonstrated a good differentiation with an AUC of 0.92, while EMG-DUET performed worse than random chance, with an AUC of 0.42. Similarly, against the AL threshold in OBS-HATLV (**Figure 3b2**), OBS-DUET continued to show strong predictive power with an AUC of 0.91, whereas EMG-DUET’s performance was comparatively weaker, with an AUC of 0.68.

EMG-DUET and OBS-DUET exhibited distinct performance patterns when classifying task risk levels against EMG-Peak and OBS-HAL (**Table 3**). EMG-DUET achieved optimal performance at a threshold of 70%, yielding a high sensitivity of 88% and specificity of 87% in comparison to EMG-Peak. However, its performance declined when evaluated against the TLV threshold in OBS-HATLV at an optimal threshold of 60%, where specificity dropped to 17%, albeit with perfect sensitivity (100%). Against the AL threshold in OBS-HATLV, EMG-DUET reached a sensitivity of 83% and specificity of 48% at its optimal threshold of 63%.

**Table 3.** Sensitivity and specificity of EMG-DUET and OBS-DUET at their optimal decision thresholds (expressed as %) against the two reference tools, EMG-Peak and the TLV and AL in OBS-HATLV.

| Ground truth | Predictor | Best threshold | Sensitivity | Specificity |
| --- | --- | --- | --- | --- |
| EMG-Peak | EMG-DUET | 70% | 88% | 87% |
|  | OBS-DUET | 46% | 75% | 63% |
| OBS-HATLV-TLV | EMG-DUET | 60% | 100% | 16% |
|  | OBS-DUET | 52% | 100% | 79% |
| OBS-HATLV-AL | EMG-DUET | 63% | 83% | 48% |
|  | OBS-DUET | 45% | 79% | 94% |

In comparison, OBS-DUET demonstrated superior performance relative to the TLV and AL thresholds in OBS-HATLV. At an optimal threshold of 52%, it achieved a perfect sensitivity of 100% and specificity of 79% against the TLV threshold, and at 45%, it showed 79% sensitivity and 94% specificity against the AL threshold. However, OBS-DUET’s sensitivity and specificity were less robust against EMG-Peak, peaking at 75% and 63%, respectively, at an optimal threshold of 46%.

### 3.3 Correlations with occupational-level prevalence of DUE symptoms

When comparing the ranks of risk assessment outcomes to the ranks of real-world work-related symptom prevalence in the fingers, hands, and wrists across occupations, the tools demonstrated varying degrees of alignment. Although none of the correlations reached statistical significance, EMG-DUET showed a weak correlation (ρ [CIs] = 0.35 [-0.23, 0.78], p = 0.215; **Figure 4a**), and EMG-Peak exhibited a moderate correlation (ρ = 0.46 [-0.16, 0.80], p = 0.100; **Figure 4c**). In contrast, OBS-DUET (ρ = 0.00 [-0.61, 0.59], p = 0.994; **Figure 4b**) and OBS-HATLV (ρ = 0.09 [-0.56, 0.67], p = 0.764; **Figure 4d**) showed no meaningful correlation with prevalence data.

**Figure 4.**
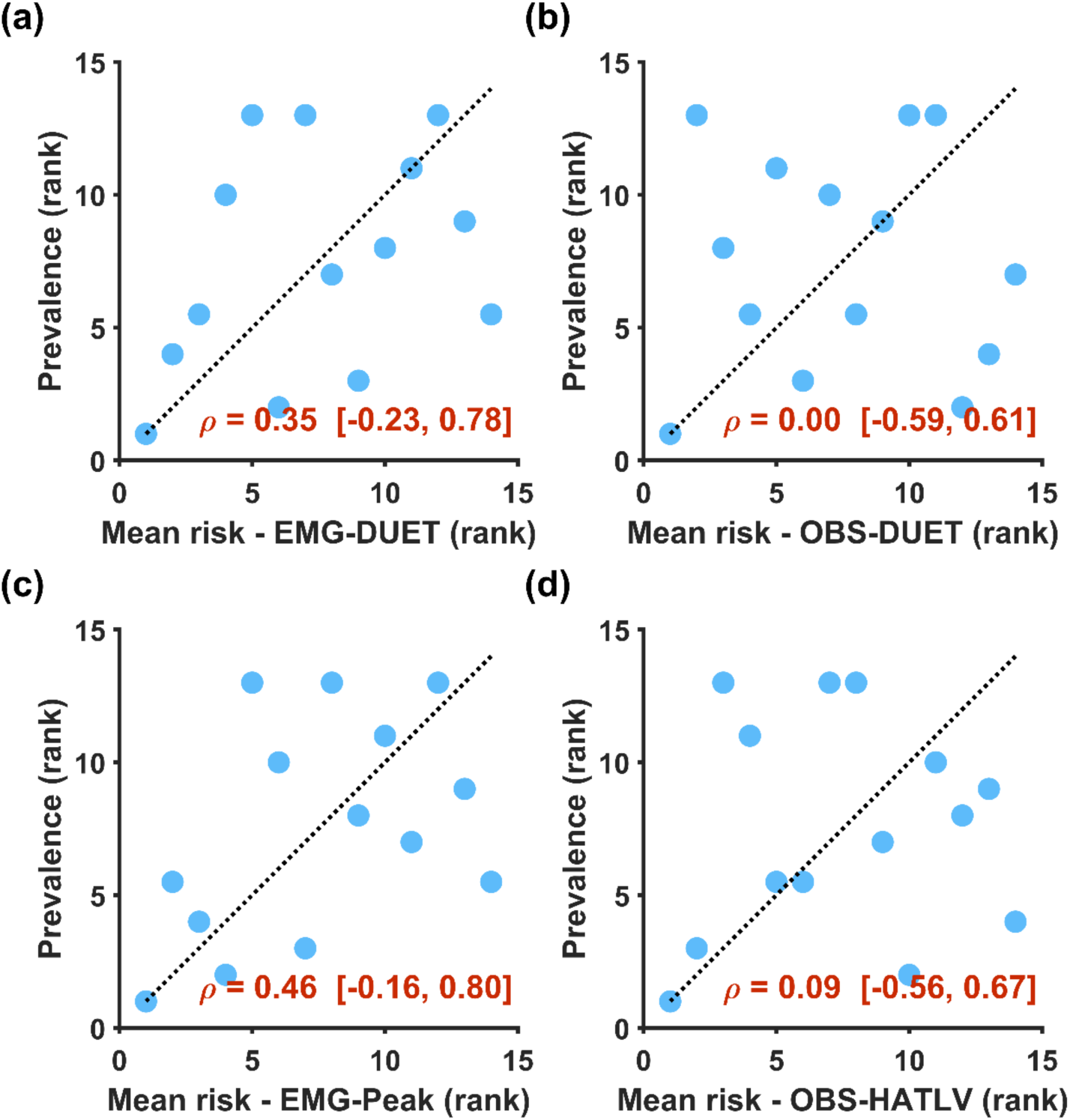
Rank comparisons between aggregated occupation-level risk assessment outcomes from (a) EMG-DUET, (b) OBS-DUET, (c) EMG-Peak, and (d) OBS-HATLV, and the corresponding occupational-level prevalence of DUE symptoms in the fingers, hands, and wrists. Each point represents an occupation. Spearman’s correlation coefficients (ρ) are shown in each panel.

## 4 Discussion

As a proof of concept, this study explored whether an EMG-based implementation of DUET (EMG-DUET) could provide meaningful risk rankings for DUE symptoms in hand-intensive work. Regarding input modality, tools using the same modality aligned more closely with each other than EMG-based and observation-based tools, suggesting that the two approaches capture overlapping but partly distinct aspects of physical exposure. Consistent with this pattern, EMG-based tools showed numerically stronger, although non-significant, correlations with occupational-level DUE symptom prevalence than observation-based tools. Regarding modelling framework, both DUET variants aligned most closely with their corresponding non-DUET benchmarks; however, DUET-based tools showed weaker correlations with prevalence than the respective non-DUET tools.

### 4.1 DUET versus non-DUET tools

Both DUET-based tools aligned most closely with non-DUET benchmark tools using the same input modality. EMG-DUET aligned with EMG-Peak, as shown by a moderate rank correlation with a value closer to the strong than the weak threshold (r_rm_ = 0.63; **Figure 2c**) and high classification performance against the EMG-Peak threshold (AUC = 0.94; sensitivity = 88% and specificity = 87% at the optimal threshold; **Figure 3a**, **Table 3**). This alignment was expected because both tools were derived from normalized forearm muscle activity (%MVE), although EMG-DUET further transformed this input through the DUET fatigue-failure framework.

Similarly, OBS-DUET aligned with OBS-HATLV, showing a strong rank correlation (r_rm_ = 0.72; **Figure 2f**) and high classification performance against both the TLV (AUC = 0.92) and AL (AUC = 0.91) thresholds (**Figure 3b1,b2**). This finding is consistent with previous work showing strong associations between OBS-DUET and other observation-based ergonomic risk assessment tools, such as the Revised Strain Index (Jorgensen et al., 2024).

Although DUET-based and non-DUET tools using the same input modality showed clear alignment, they are not conceptually equivalent. For example, both OBS-DUET and OBS-HATLV use exertion intensity and repetition as core inputs, but they integrate these inputs differently. OBS-HATLV combines hand activity and normalized peak force into a threshold-based score, whereas DUET translates exertion intensity and repetition into cumulative damage through a fatigue-failure framework. Therefore, strong alignment between OBS-DUET and OBS-HATLV supports consistency in risk ranking, but does not imply that the two tools capture risk through the same computational pathway.

These findings suggest that the DUET framework can retain input-specific risk-ranking patterns when exertion estimates are derived from either observation or EMG. However, this alignment should not be interpreted as equivalence or validation of EMG-DUET as a standalone risk assessment tool. Rather, it indicates that inserting EMG-derived exertion estimates into the DUET framework produced risk rankings that were broadly consistent with an EMG-based benchmark, while the original observation-based DUET remained consistent with an observation-based benchmark.

### 4.2 EMG-based versus observation-based tools

Although EMG-Peak and OBS-HATLV are both intended to reflect risk-relevant exposures of the DUE, they showed only a moderate rank correlation across the same tasks, with a value closer to the weak than the strong threshold (r_rm_ = 0.41; **Figure 2b**). A similar pattern was observed between the two DUET variants, where EMG-DUET and OBS-DUET also showed a moderate correlation closer to the weak than the strong threshold (r_rm_ = 0.42; **Figure 2a**). These findings suggest that EMG-based and observation-based tools are not interchangeable, even when applied to the same hand-intensive tasks.

This divergence may reflect differences in the aspects of physical exposure captured by the two input modalities. sEMG provides a physiological measure of forearm muscle activation and may reflect force-generating contractions, stabilizing activity, and co-contraction that are not directly visible during observation (Batista et al., 2024; Kimura et al., 2007). However, EMG signals are also sensitive to electrode placement, perspiration, movement artefacts, posture, and task-specific EMG–force relationships, all of which can introduce physiological and measurement variability (Abdoli-Eramaki et al., 2012; Clancy et al., 2002). In contrast, observation-based tools estimate exertion intensity, repetition, and duty cycle from visible task characteristics, making them more directly linked to observable work patterns but also susceptible to inter- and intra-rater variability and systematic observer bias (Dahlgren et al., 2022; Nyman et al., 2023).

These differences help explain why the strongest associations occurred within the same input modality, while associations across EMG-based and observation-based tools were weaker. EMG-based tools may capture muscular loading that is difficult to infer visually, whereas observation-based tools may better capture contextual features such as hand activity, movement frequency, and visible task organization. Therefore, the two approaches should be regarded as complementary rather than interchangeable sources of exposure information.

### 4.3 Alignment with occupational-level prevalence

To explore the potential correspondence between tool-derived risk rankings and occupational health outcomes, aggregated occupation-level risk rankings were compared with prevalence estimates of work-related symptoms in the fingers, hands, and wrists. Because task-level risk estimates were averaged within participants and then aggregated to match occupational-level prevalence data, the analysis could not preserve differences among tasks, workers, schedules, and recovery patterns within the same occupation. The comparison should therefore be interpreted as an exploratory assessment of occupation-level alignment rather than a validation of predictive capacity.

Within this exploratory context, although none of the correlations reached statistical significance, two distinct patterns emerged. First, EMG-based tools showed numerically stronger correlations with the prevalence proxy than observation-based tools. EMG-DUET showed a weak correlation (ρ = 0.35, p = 0.215), and EMG-Peak showed a moderate correlation closer to the weak than the strong threshold (ρ = 0.46, p = 0.100), whereas OBS-DUET (ρ = 0.00, p = 0.994) and OBS-HATLV (ρ = 0.09, p = 0.764) showed negligible correlations (**Figure 4**). Second, within each input modality, DUET-based tools showed weaker correlations with the prevalence proxy than their corresponding non-DUET benchmarks. These patterns suggest that both input modality and modelling framework may influence how task-level exposure rankings correspond to occupation-level symptom prevalence.

The numerically stronger correlations observed for EMG-based tools are partly consistent with previous evidence linking peak forearm muscular load to discomfort and diagnosed DUE symptoms (Nordander et al., 2013). However, this pattern should not be interpreted as evidence that EMG-based tools are superior to observation-based tools. Previous studies have reported limited correlations between EMG-Peak and DUE prevalence, whereas observation-based HATLV has shown predictive validity for outcomes such as carpal tunnel syndrome (Bonfiglioli et al., 2013; Dahlqvist et al., 2024; Yung et al., 2019). The present pattern may instead reflect differences in the exposure characteristics captured by each input modality, together with the limitations of comparing task-level measurements with aggregated occupational prevalence.

The weaker correlations observed for DUET-based tools also require cautious interpretation. DUET transforms exertion intensity and repetition into cumulative damage through a nonlinear fatigue-failure framework, whereas EMG-Peak and OBS-HATLV retain more direct representations of peak muscular load or hand activity and force. The current DUET implementation may therefore transform or compress task-level exposure differences in ways that reduce their correspondence with occupational-level prevalence estimates. This finding does not invalidate the DUET framework, but it indicates that further calibration of its modelling steps may be needed, particularly when EMG-derived exertion estimates are used as input.

Future studies using real-world and full-day exposure data and prospective symptom follow-up are needed to determine whether EMG-DUET, OBS-DUET, or combined EMG-observation approaches can predict DUE symptom risk in real-world occupational settings.

### 4.4 Practical Implications

Modern workplaces often involve varied tasks, fluctuating workloads, and diverse work schedules. In such settings, ergonomic risk assessment tools need to be both flexible enough to integrate exposures across tasks and interpretable enough to support practical decision-making. The DUET framework has a practical advantage in this regard because it estimates cumulative damage at the task level, which can then be integrated or weighted across task rotations, workers, or work schedules (Mehdizadeh et al., 2020). Its probabilistic output may also support communication with stakeholders by translating complex exposure patterns into an interpretable estimate of symptom risk (Gallagher et al., 2018; Mehdizadeh et al., 2020).

The present findings suggest that EMG-derived exertion estimates can be inserted into this framework while retaining risk-ranking patterns broadly consistent with an EMG-based benchmark. This supports the potential value of EMG-DUET as a sensor-informed extension of cumulative ergonomic assessment, particularly for hand-intensive work where continuous or repeated visual assessment may be resource-intensive. At the same time, the weaker alignment between EMG-based and observation-based tools indicates that EMG should not be viewed simply as a replacement for observation. Instead, EMG and observation may provide complementary information: EMG can capture physiological muscle activation, whereas observation can capture visible task organization, hand activity, and contextual features of work.

Therefore, a practical implication of this study is that future ergonomic assessment may benefit from hybrid strategies rather than relying on a single measurement approach. Observation-based tools may remain useful for rapid screening and contextual interpretation, whereas EMG-based approaches may be valuable when more detailed or continuous information on muscular loading is needed. Because the EMG-DUET framework is modular, future improvements in wearable EMG technology, signal processing, and automated cycle detection could be incorporated into the same cumulative-risk structure. However, until further calibration and validation are available, EMG-DUET should be regarded as an exploratory tool for method development rather than a standalone tool for routine risk assessment.

### 4.5 Limitations

As an exploratory study, this work has several methodological and technical limitations that should be considered when interpreting the findings. First, muscle activity and observed exertion were recorded during brief task segments, with an average duration of 7.3 minutes. Because full-day work schedules were not available, task-level risk estimates were projected to a standardized six-hour exposure period to enable comparison across tools. Although this standardization allowed task rankings to be compared within a common analytical framework, it does not capture the variability of real workdays, including task rotation, work pace, rest breaks, and recovery periods. Future studies should therefore use full-shift recordings and incorporate real-time work-rest cycles.

Second, the comparison with occupational-level prevalence estimates relied on several simplifications. Task-level risk estimates were averaged and aggregated to match occupational categories, which may have obscured differences among tasks, workers, schedules, and exposure distributions within the same occupation. In addition, the prevalence data represented symptoms in the fingers, hands, or wrists at the occupational level, rather than individual-level prospective health outcomes. Therefore, the prevalence analysis should be interpreted as a preliminary external comparison, not as validation of predictive capacity.

Third, the EMG-based pipeline relied on assumptions that require further calibration. Through-forearm sEMG provides a practical composite signal from the forearm flexor–extensor region, but it is not equivalent to muscle-specific EMG and remains sensitive to electrode placement, posture, movement artefacts, perspiration, and task-specific EMG–force relationships. Similarly, the conversion from normalized muscle activity to estimated exertion was based on previously published modelling work and was not optimized for all task types, postures, contraction modes, or individuals in the present dataset. These assumptions may influence cycle identification, exertion estimates, and final risk scores. Future work should therefore include sensitivity analyses and empirical calibration of the EMG-to-exertion model against biomechanical or force-based reference measures.

Fourth, EMG-Peak was calculated from through-forearm EMG rather than from the traditional extensor-specific placement used in the proposed action level for forearm muscular load. Because no established action level exists for through-forearm EMG, the 30% MVE threshold was used as an approximate benchmark for comparative purposes, not as a validated threshold for this electrode configuration. This choice may have affected risk classification and correlations with EMG-DUET.

Finally, the DUET framework itself involves modelling assumptions that may limit generalizability. The fatigue-failure model maps exertion to cumulative tissue damage using simplified relationships derived from prior biomechanical and epidemiological work. It does not explicitly account for inter-muscle coordination, antagonistic co-activation, tissue adaptation, or recovery during rest breaks. In the present study, DUET-based scores also appeared to compress the range of risk estimates, particularly for EMG-DUET, which may reduce sensitivity to differences among tasks. Addressing these limitations will require targeted refinement of signal processing, cycle identification, exertion-to-damage mapping, and validation against full-shift exposure data with prospective DUE symptom follow-up.

## 5 Conclusions

This proof-of-concept study explored whether an EMG-based implementation of DUET (EMG-DUET) could provide meaningful risk rankings for DUE symptoms in hand-intensive work. Regarding input modality, EMG-based and observation-based tools showed weaker alignment with each other than with tools using the same input modality, suggesting that they capture partly distinct aspects of physical exposure. Consistent with this distinction, EMG-based tools also showed numerically stronger, although non-significant, correlations with occupational-level DUE symptom prevalence than observation-based tools. Regarding modelling framework, both DUET variants aligned most closely with their corresponding non-DUET benchmarks; however, DUET-based tools showed weaker correlations with prevalence than the respective non-DUET tools. These findings suggest that risk rankings and their correspondence with occupational-level symptom prevalence differ according to both input modality and modelling framework. EMG-DUET should therefore be regarded as an exploratory implementation rather than a validated standalone risk assessment tool. Future work should refine the EMG-to-exertion conversion, cycle identification, and damage mapping, and evaluate the approach using full-shift exposure data collected under real-world conditions and prospective DUE symptom follow-up.

## Data Availability

All data produced in the present study are available upon reasonable request to the authors

## Acknowledgement

We thank Eric Martini Linger for assistance with data collection and Professor Emeritus Sean Gallagher from Samuel Ginn College of Engineering at Auburn University for valuable scientific and technical guidance. We are also grateful to the participating companies and organizations for their support with recruitment and for providing measurement locations. Finally, we extend our sincere appreciation to all participants for their time and effort.

## CRediT authorship contribution statement

**Xuelong Fan:** Conceptualization, Methodology, Formal analysis, Software, Data Curation, Investigation, Writing - Original Draft, Writing - Review & Editing, Visualization, Project administration; **Johan Rydgård:** Software, Data collection, Data Curation, Writing - Review & Editing; **Pasan Hettiarachchi:** Methodology, Writing - Review & Editing; **Kristina Eliasson:** Data collection, Writing - Review & Editing; **Camilla Dahlqvist:** Data collection, Writing - Review & Editing; **Peter J. Johansson:** Conceptualization, Methodology, Data collection, Resources, Project administration, Writing - Review & Editing, Supervision, Funding acquisition.

## Funding sources

This work was supported by AFA Insurance [Ref. No. 200070].

## Declaration of competing interest

There are no conflicts of interest to declare.

## Declaration of generative AI and AI-assisted technologies in the writing process

During the preparation of this work the author(s) used ChatGPT (GPT-5.5) in order to improve the language and readability. After using this tool/service, the author(s) reviewed and edited the content as needed and take(s) full responsibility for the content of the publication.

## Supplement

**Table S1.** The names and descriptions of tasks used in this study (N = 78). The task ID is composed as “sex” + “participant ID” + “side letter” + “-”+ “occupation abbreviation” + “task number”. F and M refer to female and male, respectively. Side letter L stands for the left side, without which it refers to the right side.

| Task ID | Occupation | Task description | Task ID | Occupation | Task description |
| --- | --- | --- | --- | --- | --- |
| **F12-DTW01** | Dental technician work | Grinding a dental prosthesis. | **F12-DTW02** | Dental technician work | Wax-up of a dental prosthesis. |
| **F12-DTW03** | Dental technician work | Bonding and casting of a dental prosthesis. | **F12-DTW04** | Dental technician work | Devesting the mold with a small hammer. |
| **F12-DTW05** | Dental technician work | Administrative work. | **F12-DTW06** | Dental technician work | Packaging of the product. |
| **F12-DTW07** | Dental technician work | Bend wire. | **F12-DTW08** | Dental technician work | Polish metal. |
| **F12-DTW09** | Dental technician work | CAD work on a computer. | **F12-DTW10** | Dental technician work | Applying thermoset plastic. |
| **F12L-DTW01** | Dental technician work | Grinding a dental prosthesis. | **F12L-DTW02** | Dental technician work | Wax-up of a dental prosthesis. |
| **F12L-DTW03** | Dental technician work | Bonding and casting of a dental prosthesis. | **F12L-DTW07** | Dental technician work | Bend wire. |
| **F12L-DTW08** | Dental technician work | Polish metal. | **F13-PW01** | Painting work | Washing the exterior wall before painting. |
| **F13-PW02** | Painting work | Scraping the wall prior to painting. | **F13-PW03** | Painting work | Outdoor wall painting. |
| **F13-PW04** | Painting work | Outdoor painting in hard-to-reach areas. | **F13-PW05** | Painting work | Carrying a painter's step stool. |
| **F13-PW06** | Painting work | Painting from a mobile elevated work platform. | **F14-EW01** | E-warehouse work | Sorting of products (big bags). |
| **F14-EW02** | E-warehouse work | Sorting of products (shoes). | **F14-EW03** | E-warehouse work | Pull a pallet by hand. |
| **F18-M01** | Massage | Massage. | **F19L-HC01** | Hotel cleaning | Make a bed in a hotel. |
| **F19L-HC02** | Hotel cleaning | Clean a toilet in a hotel. | **F19L-HC03** | Hotel cleaning | Mopping the floors in a hotel. |
| **M04-MW01** | Manufacturing work | Installation and sealing of prefabricated sink units. | **M04-MW02** | Manufacturing work | Installation of prefabricated kitchen cabinets onto modular building elements. |
| **M04-PW03** | Painting work | Painting the wall with a roller. | **M05-VA01** | Vehicle assembly | Vehicle assembly on an assembly line - Task 1. |
| **M05-VA02** | Vehicle assembly | Vehicle assembly on an assembly line - Task 2. | **M05-VA03** | Vehicle assembly | Vehicle assembly on an assembly line - Task 3. |
| **M05-VA04** | Vehicle assembly | Vehicle assembly on an assembly line - Task 4. | **M05-VA05** | Vehicle assembly | Vehicle assembly on an assembly line - Task 5. |
| **M05-VA06** | Vehicle assembly | Vehicle assembly on an assembly line - Task 6. | **M05-VA07** | Vehicle assembly | Vehicle assembly on an assembly line - Task 7. |
| **M05-VA08** | Vehicle assembly | Vehicle assembly on an assembly line - Task 8. | **M06-EA01** | Electronic assembly | Cable assembly. |
| **M06-EA02** | Electronic assembly | Cut the shield on cables with pliers. | **M06-EA03** | Electronic assembly | Crimp the end sleeve on a cable with new pliers. |
| **M06-EA04** | Electronic assembly | Crimp the end sleeve on the cable with old pliers. | **M06-EA05** | Electronic assembly | Spiralizing cable with the old method. |
| **M06-EA06** | Electronic assembly | Spiralizing cable with a new method. | **M06-EA07** | Electronic assembly | Turning off the cable with the new tool. |
| **M06-EA08** | Electronic assembly | Turning of cable with an old tool. | **M06-EA09** | Electronic assembly | Applying tubing to the cable. |
| **M06-EA10** | Electronic assembly | Crimping lugs on cable with a hydraulic crimper. | **M07-CW01** | Computer work | Computer work - Task 1. |
| **M08-CW01** | Computer work | Computer work - Task 2. | **M09-WR01** | Window reglazing | Installing glass in windows. |
| **M09-WR02** | Window reglazing | Cleaning window putty with a hammer and knife. | **M09-WR03** | Window reglazing | Cleaning window putty with a vibrating cleaning machine. |
| **M09-WR04** | Window reglazing | Carrying and cutting glass. | **M09-WR05** | Window reglazing | Apply window putty. |
| **M09-WR06** | Window reglazing | Lifting and carrying windows. | **M09-WR08** | Window reglazing | Cleaning workshop and sweeping floors with a brush. |
| **M11-CP01** | Concrete production | Leveling concrete with trowels and hand tools. | **M11-CP02** | Concrete production | Roll the concrete surface. |
| **M11-CP03** | Concrete production | Tying reinforcement mesh. | **M11-CP04** | Concrete production | Bending reinforcement mesh. |
| **M15-EW01** | E-warehouse work | Packaging of products in an e-commerce warehouse. | **M15-EW02** | E-warehouse work | Packaging of products in an e-commerce warehouse. |
| **M15-EW03** | E-warehouse work | Pick orders in an e-commerce warehouse. | **M16L-RW01** | Restaurant work | Prepare the table for dinner in a restaurant. |
| **M16L-RW02** | Restaurant work | Clear the tables in a restaurant. | **M17-RW01** | Restaurant work | Cut and chop vegetables. |
| **M17-RW02** | Restaurant work | Stir with a large ladle in a big pot. | **M17-RW03** | Restaurant work | Cut and chop tomatoes. |
| **M20-FC01** | Fine carpenter | Assemble Wooden Tables. | **M20-FC02** | Fine carpenter | Pack 120 tables in a carton. |
| **M21-MW01** | Metal work | Loading parts into a welding fixture. | **M22-MW01** | Manufacturing work | Mounting of cable ladder on the wall - Task 1. |
| **M22-MW02** | Manufacturing work | Connection of an appliance box. | **M23-MW01** | Manufacturing work | Mounting of cable ladder on the wall - Task 2. |
| **M23-MW02** | Manufacturing work | Connection of an appliance box. | **M23-MW03** | Manufacturing work | Pick products on the shelves. |

## Notes

### Competing Interest Statement

The authors have declared no competing interest.

### Author Declarations

Ethics committee/IRB of the Swedish Ethical Review Authority in Gothenburg gave ethical approval for this work

### Summary of Updates

We changed the title and did major edition regarding how the data was presented. Notably, no results were changed.

